# Prevalence and associated risk factors of preterm and post-term births in Northern Ghana: a retrospective study in Savelugu Municipality

**DOI:** 10.1101/2023.10.21.23297356

**Authors:** Silas Adjei-Gyamfi, Abigail Asirifi, Hirotsugu Aiga

## Abstract

**Introduction:** Preterm and post-term births are prominent leading causes of neonatal mortalities and significant contributors to long-term adverse health outcomes. Although preterm and post-term births are disproportionately rampant in most parts of Ghana, the magnitude and underlying predictors are not well comprehended which necessitates more evidence for appropriate interventions. This study assessed the prevalence and identified vital risk factors of preterm and post-term births in Northern Ghana.

**Methods:** This study is a retrospective cross-sectional design conducted on 356 postnatal mothers from February to March 2022 in Savelugu Municipality of Northern Region, Ghana. Anthropometric, clinical, obstetric, and sociodemographic data were collected from antenatal records using structured questionnaires. Multinomial logistic regression was used to identify independent factors of preterm and post-term births at 95% confidence interval.

**Results:** Prevalence of preterm and post-term births were 19.4% and 6.5% respectively. Anaemia in the first trimester of pregnancy (AOR: 2.205; 95%CI: 1.011− 4.809), non-use of insecticide-treated bed nets (ITNs) during pregnancy (AOR:1.979; 95%CI: 0.999 − 3.920), maternal age less than 20 years (AOR:12.95; 95%CI: 2.977 − 56.34), and mothers with junior high school education (AOR: 0.225; 95%CI: 0.065 − 0.797) were independently associated with preterm births.

Predictors for post-term births were macrosomic (large birthweight) delivery (AOR:8.128; 95%CI: 1.777 − 37.18) and mothers with senior high school education (AOR:0.001; 95%CI: 0.0001 − 0.125).

**Conclusion:** Preterm births are very prevalent, while post-term births are increasingly becoming crucial in the municipality. These nutritional (gestational anaemia) and non-nutritional (ITNs use, teenagers, maternal education, and macrosomic births) predictors of preterm and post-term deliveries are modifiable and preventable. Therefore, interventions should be targeted at intensified community education on nutrition and lifestyle modifications, in addition to vigorous promotion of girls’ child education through parental empowerment.

**KEY MESSAGES:**

- Globally, most studies do not consider a wider spectrum of variables to identify risk factors of preterm and post-term births while few researchers consider studies on post-term births. Although preterm and post-term births are disproportionately common in some parts of Ghana, the magnitude and underlying determinants are not well understood which requires more evidence for suitable interventions in Northern Region and Ghana at large.
- We found the prevalence of preterm and post-term births to be relatively high in the Savelugu municipal. Prominent nutritional and non-nutritional factors were identified to be responsible for the delivery of preterm and post-term babies.
- To achieve better pregnancy outcomes, the promotion of girl child education through parental empowerment, rigorous community nutrition education, and lifestyle modification are recommended.

## INTRODUCTION

Preterm and post-term births are regarded as the two forms of inappropriate or abnormal gestational age at delivery responsible for chronic maladies and mortalities worldwide ^(1,2)^. Preterm births have predominantly been established as the global principal cause of mortalities among children under five years of age accounting for 35.0% of deaths among neonates and about 16.0% of all deaths worldwide ^(3,4)^. Similarly, post-term births are associated with long-term child morbidities and mortalities ^(5,6)^. The World Health Organization denotes preterm deliveries (also known as prematurity) as births that occur before 37 gestational weeks whilst post-term births (also called post-maturity/post-date) are defined as deliveries at a gestational age equal to or above 42 weeks ^(4)^.

The multi-factorial determinants of abnormal gestational age at birth are largely related to biological, psycho-emotional, obstetric, economic, demographic, and social contributory factors ^(1,6)^. Some significant elements associated with preterm births encompass antepartum haemorrhage, pre-eclampsia, multiple pregnancy ^(7–11)^, gestational anaemia, rural residence, short birth spacing ^(1,12–15^), multiparity, adolescent mothers, and low socioeconomic status (SES) ^(11,16)^, married mothers, low parity, and inadequate antenatal visits ^(16,17)^. Asynchronously, other studies revealed large babies ^(18)^, maternal overweight, low gestational weight gain ^(19,20)^, high SES, unmarried mothers ^(5)^, pregnancy-related psycho-emotional problems ^(21)^ as predictors of post-term infants. This highlights the need to investigate some nutritional-related predictors of these preterm and post-term deliveries.

The global action for preterm births particularly centered on policies including the Sustainable Development Goals (SDGs) and Global Strategy for Women’s, Children’s and Adolescents’ Health is still on course to improve newborn and child survival. However, post-dates have not been accorded similar needed attention. Notwithstanding, the prevalence rates of preterm and post-term births are still on the rise as preterm births have significantly upsurged spanning over 20 years ^(22)^. Globally, an estimated 13.4 million (9.9%) babies were born too soon in 2020 with the highest burden in Southern Asia (13.2%) and sub-Saharan Africa (10.1%) ^(4)^. The prevalence of preterm deliveries in Brazil ^(16)^, Ethiopia ^(9,12)^, Nigeria ^(11)^, Kenya ^(10)^, and Southern Ghana ^(17)^ were 11.5%, 13.3%, 16.0%, 18.3%, and 18.9% respectively. Additionally, in the Kassena-Nankana district in the Upper East Region of Ghana, the prevalence of preterm births was 32.0%^(23)^. On the other hand, despite data paucity on post-matured births in Ghana, the incidence was found to be 4.5%, 6.0%, 6.5%, and 11.4% among the American ^(20)^, Ethiopian ^(5)^, British ^(24)^, and South African ^(25)^ mothers correspondingly in some prospective and retrospective reports.

The entire globe has unacceptable gaps in perinatal outcomes by race and for the poorest and most marginalized groups in society. Nonetheless, preterm and post-date births have higher survival in developed nations after delivery while low-resource settings like Ghana are still recording huge survival gaps ^(1,4)^. Therefore, pinpointing and comprehending the risk factors for preterm and post-term births have the potential to help tackle this problem and also aid in achieving SDG 3 through the reduction of perinatal and neonatal mortalities and morbidities in developing countries including Ghana. Globally, most maternal and child health information on risk factors for abnormal gestational age at delivery are always limited to prematurity without paying much attention to post-maturity. Nevertheless, this study will consider both. Although preterm and post-term births are disproportionately rampant in most parts and peri-urban belts of Northern Ghana, the magnitude and underlying predictors are also not well comprehended which necessitates more evidence for apt interventions. Currently, there is a scarcity of published data on the prevalence of abnormal gestational age at delivery and its possible determining factors in the Savelugu municipality and Northern Ghana. This study therefore assessed the prevalence of preterm and post-term births and their associated risk factors among mothers attending postnatal care services in the Savelugu municipality of Northern Ghana.

## MATERIALS AND METHODS

### Study design

The study employed a retrospective cross-sectional design to determine the magnitude of preterm and post-term deliveries among mothers attending postnatal care (PNC) services and also identify the predictors of these adverse pregnancy outcomes in the study area.

### Study area

Savelugu municipality is situated in the northern hemisphere of Northern Region of Ghana. The major source of income in the municipal is farming while the inhabitants are predominantly from the Dagomba ethnic group. Five major public health facilities with 21 operational CHPS zones provide health services to a populace of 125,469 in this municipality. The total number of reproductive-aged women is more than 40,000 which places the municipal at a higher risk of frequent pregnancies with routine increased rate of PNC and antenatal care (ANC) attendances. The municipal has a skilled delivery rate of 93% whereas monthly skilled delivery is estimated between 300 to 400. Moreover, maternal and child health record books (MCHRBs) have been consistently used by mothers when seeking antenatal, postnatal, and child welfare services since the inception of a Japanese project on MCHRBs ^(26)^.

### Study population

This study recruited 356 postnatal mothers having a baby of less than 29 days (neonatal period) and living in the municipality. Mothers who had MCHRBs and attending first-day PNC services at the health facilities were sampled while those with heart diseases, multiple births, and home deliveries were not included in the study.

### Sample size and sampling methods

The sample size was attained by the formula; 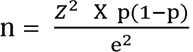. Due to the scarcity of Ghanaian data on post-term births, the study employed prevalence (p) of preterm births in Southern Ghana which was reported as 33% ^(8)^. By applying margin of error (e) of 5%, and standard normal variate (z) at 95% confidence level of 1.96, the sample size was initially estimated at 339. The final sample size was rounded off to 356 after adding 5.0% non-response rate ^(7)^. Afterward, probability proportional to size technique was used to estimate the sample sizes for the five major public health facilities in the study area (Table 1). A coin was then flipped to randomly select all the 356 postnatal mothers, using the daily PNC registry from the respective health facilities.

### Study variables

All the study variables and/or data were collected from MCHRBs and/or ANC records except some sociodemographic information which was collected through structured interviews in February and March 2022. Data collection was done using structured questionnaires designed onto Open Data Kit (ODK) version 2021.2.4 (Get ODK Inc., San Diego, USA), and pre-installed onto handheld tablets.

The outcome variable for this study is gestational age at delivery which is multinomial recorded as preterm (< 37 weeks) = 1, normal term (≥ 37–41 weeks) = 2, and post-term (≥ 42 weeks) = 3 where the “normal term” was used as the base outcome (reference point) during analyses.

From the collected data, 39 exposure variables were selected and categorized based on biological plausibility, existing literature, and the potential to influence gestational age at delivery ^(18,27,28,29)^ (Tables 2, 3, and 4). Some of these exposure variables include marital status, educational level, ethnic group, religion, sex of neonate, household size, job status, birthweight, birth length, parity, gravidity, frequency of ANC visits, tetanus-diphtheria immunization, iron-folic acid intake, anthelminthic drugs intake, and sulphadoxine-pyrimethamine (SP) doses intake, gestational weight gain (measured within one week prior to delivery and the one recorded at the first trimester ANC visit), pre-gestational body mass index (mother’s weight at first trimester ANC visit used as a proxy for pre-pregnancy weight since foetal weight is low). Other variables were diagnosis of hypertension, diabetes, hepatitis B, and malaria during pregnancy, rhesus type, Glucose-6-phosphate dehydrogenase deficiency (G6PD) status, haemoglobin levels at first, second, and third pregnancy trimesters (classified as anaemia [< 11g/dL], normal [≥ 11– 13.1g/dL], and high [≥ 13.2g/dL]), and wealth index ^(18,27,28)^.

Wealth index was assessed based on possession of household assets, housing quality, and availability of utilities among others which was employed as a proxy indicator for SES of mothers. By using principal component analysis, the SES of mothers was then categorized into five groups (thus, wealth quintiles: poorest, poorer, poor, less poor, least poor) ^(29)^.

### Statistical analysis

The data attained was first transferred from ODK platform onto Microsoft excel version 16.6 before moving them onto STATA. All analyses were conducted in STATA version 17.0 (Stata Corporation, Texas, USA) and results were presented in the form of tables and summary statistics in odds ratios (OR), with p-values and 95 % confidence intervals (CI). Associations between each exposure variable and gestational age at delivery were explored at the bivariate level (Chi-square/Fisher’s exact tests) (Tables 2,3,4) and those significant at p < 0.05 were simultaneously entered into a multinomial logistic regression model (Table 5). Variance inflation factor (VIF) was used to address multicollinearity between the possible exposure variables before employing them in the logistic analyses. During the testing, the variables with a VIF of less than 10 were selected for the logistic analyses.

## RESULTS

### Prevalence of preterm and post-term births

This study used all 356 completed data from the recruited mothers in the Savelugu municipal for the analysis. Out of the total sampled data, the prevalence rates of preterm and post-term births were 19.4% (95%CI: 15.4%−23.8%) and 6.5% (95%CI: 4.1%−9.5%) respectively (Figure 1).

**Fig 1.**
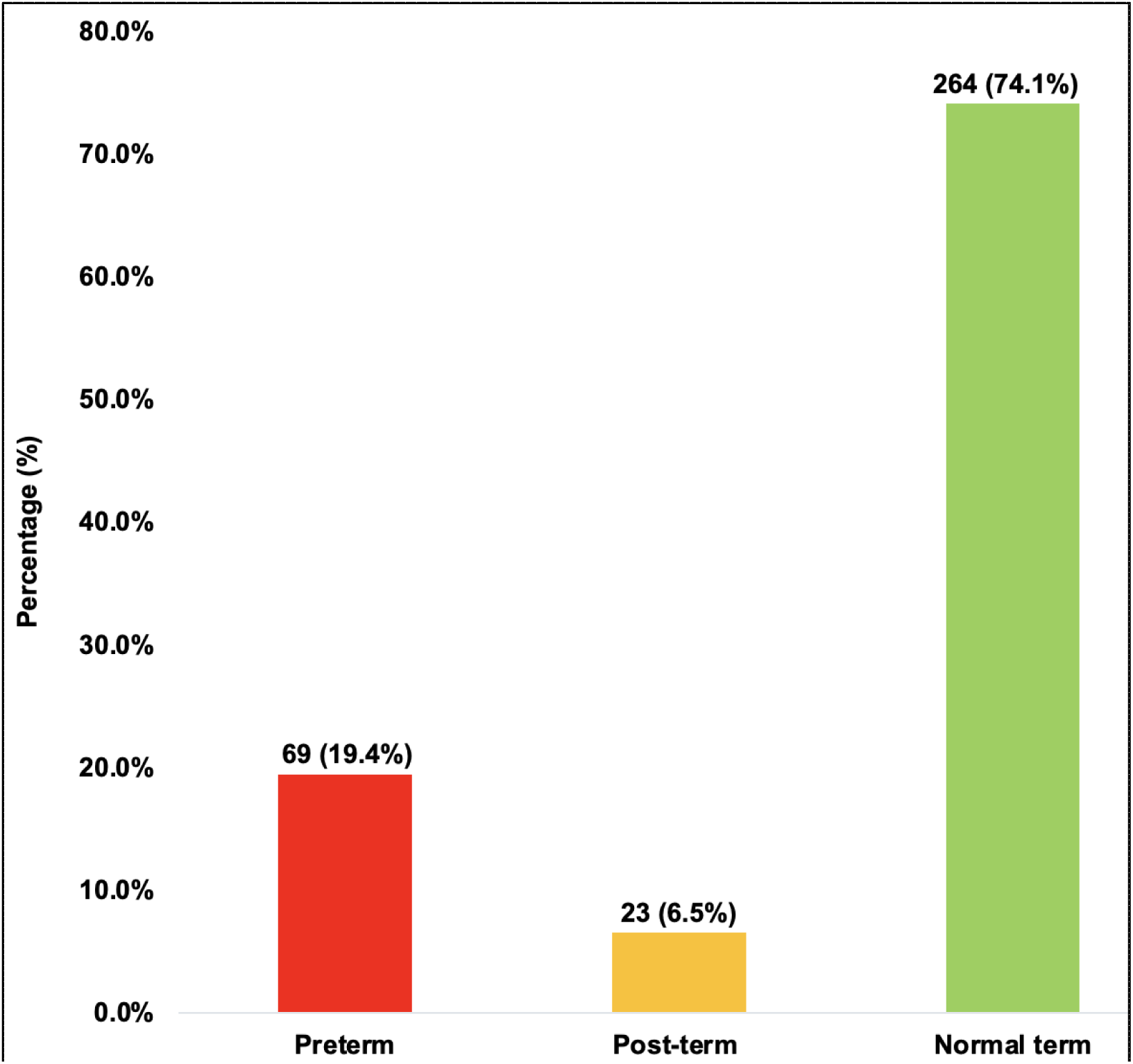
Prevalence of preterm and post-term births

### Sociodemographic and other background variables among study participants

Larger proportion of the mothers were married (90%), Muslims (88%), and aged from 20 to 35 years (87%) with a mean age of 27 years. Approximately, one-third of the respondents had no formal education. Female neonates (44%) were less than the males (56%) with about 70% of the neonates aged more than seven days of life (Table 2). Slightly above one-half of the respondents were informally employed (51%), attended ANC less than eight times (54%), took more than three doses of sulphadoxine-pyrimethamine (SP) drugs (55%), and used insecticide-treated bed nets (ITNs) (61%) during gestation (Table 3). The prevalence of gestational anaemia in the first, second, and third trimesters were 45%, 56%, and 44% respectively. Less than one-quarter of mothers were diagnosed with gestational diabetes (6%), gestational hypertension (12%), and hepatitis B (11%) infections during pregnancy. Most mothers had experienced at least two pregnancies (75%) and deliveries (73%), while a greater proportion of the neonates were born with small weight (22%) as compared to macrosomic births (9%) (Table 4).

### Association of preterm and post-term births with background variables

As presented in Tables 2 – 4, maternal age (p=0.001), educational level (p=0.009), gestational weight gain (p=0.012), birth length (p=0.041), birth weight (p<0.001), frequency of ANC visits (p<0.001), use of ITNs (p=0.037), SP intake (p=0.009), number of pregnancies (p=0.002) and deliveries (p=0.001), haemoglobin levels in each trimester of pregnancy (p<0.001), gestational diabetes (p=0.014), and G6PD status (p=0.034) showed bivariate relationship with gestational age at delivery.

### Associated risk factors of preterm and post-term births

Fifteen exposure variables exhibited significant bivariate association (p<0.05) with preterm and post-term births as shown in Tables 2 – 4. However, one variable (thus, number of pregnancies) was excluded from the significant exposure variables due to multicollinearity before all the other variables were entered into the final logistic model. In the final model, maternal age < 20 years, educational level (junior high school), non-use of ITNs, and anaemia in the first trimester of pregnancy remained significant for preterm births while senior high school level of education and macrosomia showed significant relationship with post-term deliveries (Table 5).

The result indicates that mothers aged less than 20 years (teenagers) had 12.95 (95%CI: 2.977−56.34) times increased risk of giving birth to preterm babies. Mothers who are junior high school leavers had reduced risk for preterm delivery, indicating 77.5% [=(1−0.225)×100] reduction in the delivery of preterm babies (95%CI: 0.065−0.797). Mothers who didn’t use ITNs during pregnancy had 1.979 (95%CI: 0.999−3.920) times higher risk of preterm delivery. The odds of a mother with anaemia in the first trimester of pregnancy having preterm delivery was 2.205 (95%CI: 1.011−4.809) times. Furthermore, mothers who delivered macrosomic (large weight) babies had 8.128 (95%CI: 1.777−37.18) times increased risk of post-term delivery. Mothers who completed senior high school had a decreased risk of giving birth to post-term babies by 99.9% [=(1−0.001)×100] (95%CI: 0.0001−0.125).

## DISCUSSION

Abnormal gestational age at delivery and its associated complications are one of the setbacks in reducing children under-five mortality in most developing countries like Ghana ^(1)^. The study found the prevalence of preterm and post-term births to be 19.4%. The prevalence of preterm births is greater than the worldwide incidence of 10.6% ^(30)^. Additionally, greater preterm prevalence was found in this study than the prevalence from developing countries including Brazil (11.5%) ^(16)^, Ethiopia (13.3%) ^(9,12)^, Iran (12.7%) ^(31)^, Kenya (18.3%) ^(10)^, and Southern Nigeria (16.0%) ^(11)^. This places the municipality at higher risk of neonatal deaths since preterm births are responsible for 35.0% of all neonatal mortalities and about 16.0% of global deaths ^(3)^. These aforementioned countries (Brazil, Ethiopia, Iran, Kenya, and Nigeria) might be effectively implementing the recommended interventions on premature births and may have comprehensive healthcare system for pregnant women leading to their relatively low preterm prevalence. Moreover, despite the diverse healthcare systems across those countries, the study designs, and larger sample sizes employed in those earlier studies ^(9–12,16,31)^ could be accountable for these variations. The present study finding was also lower than some retrospective studies in Northern India and a teaching hospital in Southern Ghana with prevalence of 35.0% ^(32)^ and 37.3% ^(7)^ respectively. The increased rate of multiple gestations in India and Southern Ghana could be responsible ^(7,32)^, as this may cause an overly stretched uterus which might precipitate preterm deliveries ^(12)^ as multiple gestations are established predictors of preterm births ^(30)^.

The prevalence of post-term births in this study was estimated at 6.5%. Our study’s post-term prevalence was lesser as compared to some findings in the United States of America ^(20)^, Northern Ethiopia ^(5)^, and South Africa ^(25)^ at corresponding rates of 4.5%, 6.0%, and 11.4%. The differences could be explained in terms of demographic and geographical features of the study populations, as well as the timing of these studies. Furthermore, the lesser post-term births in our study may be linked to the cultural beliefs of the Savelugu citizenry. Culturally, the people of Savelugu municipal regard post-term births as a curse. Due to this cultural belief, pregnant women usually ingest some local oxytocin (*kalgu-tim*) to increase uterine contractions whenever they reach term gestational age ^(26)^, leading to early delivery of their babies and causing a possible reduction of post-term births.

In the study, 5.1% (95%CI: 3.0% - 7.9%) of the mothers were teenagers (adolescents). Teenage mothers (thus, mothers aged less than 20 years) were 13 times more likely to deliver prematurely. An analogous pattern was found among Brazilian ^(16)^ and Nigerian ^(11)^ mothers who were less than 20 years old with an increased risk of preterm births but incongruent with some retrospective studies in Southern Ghana ^(7,17)^, Kenya ^(10)^, and Ethiopia ^(12)^. Teenagers (young mothers) are still in the active phase of physiological, psychological, emotional, biological, and physical maturity and development. These young women (teenagers) may not cope with the changing demands of pregnancy and also identify the significance of feto-maternal nutrition, childbearing, and self-care activities among others during pregnancy ^(33)^ which may lead to adverse pregnancy outcomes like preterm deliveries.

The current study demonstrated that mothers who completed junior high school had a reduced risk for preterm births. This is different from the findings of Wagura and colleagues ^(10)^ but similar to that of Spanish and Japanese cohort studies ^(34,35)^ which indicated that maternal lower (junior high school) educational level was associated with prematurity. Approximately 13.0% (95%CI: 9.6% – 16.9%) of the mothers completed junior high school while 34.0% (95%CI: 29.1% – 39.2%) had no formal education in the Savelugu municipality. Junior high school education has been linked to lower the incidence of pregnancy complications in women including delivery of preterm babies ^(36,37)^. Furthermore, it is found in most developing countries that unsatisfactory antenatal visits which predict preterm births are high in regions where junior high school enrollment rate is low ^(35,36)^. This explains why mothers who are junior high school leavers tend to be protective against premature births.

Gestational anaemia is a global challenge as well as the most prodigious haematological manifestation of pregnancy and a heralded cause of gestational complications and adverse reproductive outcomes ^(6)^. Mothers with anaemia in the first trimester of pregnancy were twice more likely to give birth to preterm babies in this study. This finding is consistent with some meta-analyses ^(38,39)^ which concluded that first-trimester anaemia increases the risk of premature births. Many prospective and retrospective studies were also consistent with our study findings ^(13–15)^ while others were incongruent ^(40,41)^. Despite the complex and multifactorial aetiological mechanisms such as nutritional deficiencies (iron, folic acid, vitamin A, or vitamin B12), infections (malaria, worm infestations, and HIV), and haemoglobinopathies (sickle cell) underlying the relationship between low gestational haemoglobin levels and prematurity, iron deficiency (which is a major contributor to gestational anaemia) tend to be the most reported trigger of premature deliveries ^(39,42)^. Thus, anaemia in the first and/or early trimester of gestation may designate pre-existing, early onset, and/or persistent iron deficiency which precipitates preterm births. Additionally, the current study revealed significant crude association between first-trimester gestational anaemia and non-intake of antihelminthics (OR: 1.55, p = 0.038). This evidence supports the mechanism of infections like worm infestations on gestational anaemia leading to the delivery of premature babies ^(42)^. Anaemia including iron deficiency could induce maternal infection, hypoxia, and oxidative stress that may trigger the spontaneous onset of preterm labour ^(32)^. This highlights the need for early prevention and treatment of anaemia during pregnancy due to the relatively high preterm prevalence in the Savelugu municipality.

About 40% (95%CI: 33.7% – 44.0%) of mothers didn’t use ITNs during pregnancy in our study which is comparable to a Nigerian cross-sectional study ^(43)^. No ITNs use during gestation was significantly associated with an increased risk of preterm births in our study. This is confirmatory to some studies in the Sahelian Africa ^(44,45)^. Strengthening community and antenatal education on the use of ITNs during pregnancy is crucial for preventing malaria infection ^(43,44)^ which predisposes pregnant women to premature deliveries. Likewise, health facilities must constantly supply these ITNs to pregnant mothers and adequately monitor ITNs use to prevent malaria infections from leading to preterm births in the municipality.

On the other hand, post-term births have not been accorded similar recognition as preterm births. From our study, senior high school leavers were protective factor for post-dates. Thus, mothers with senior high school education were less likely to deliver post-mature babies. Mothers with senior high school education were 29.5% (95%CI: 24.8% – 34.5%) in our present study. Though there are very rare studies on post-term births globally, the present finding is unparallel to reports in Ethiopia where no association was found between post-term births and maternal educational level ^(5)^. By virtue of relatively higher education among senior high school leavers, we conjecture that mothers with higher secondary education are chiefly knowledgeable and aware of their reproductive health and pregnancy outcomes and always respect antenatal education and counseling which reduces the risk of post-term births.

The prevalence of macrosomia in this study was approximately 9.0% (95%CI: 5.0% – 12.0%). Macrosomia was significantly associated with post-maturity. This finding is in contrast to that of meta-analysis ^(19)^ and longitudinal studies in the United States of America ^(20)^ but mirrored a cross-sectional study by Adjei-Gyamfi and his colleagues ^(18)^. Maternal, child, and neonatal mortalities are associated with advanced gestational age (post-term) in most developing countries^(46)^. Physiologically, large-weight (macrosomic) foetus tends to experience weekly intra-uterine weight gain estimated between 0.12 to 0.24 kilograms as the foetus continuously stays in the uterus after 37 weeks of gestation ^(47)^. As the uterus promotes growth processes, the foetus stays longer leading to late gestation. In our study, out of the 31 mothers who delivered macrosomic babies, larger proportion were diagnosed with gestational diabetes (26; 83.9%) as few mothers had no diabetes diagnosis during pregnancy (p=0.011 in χ2/Fisher’s exact test). Concurrently, of the same 31 mothers, those who had gestational weight gain (GWG) of at least 6kg (28; 90.3%) were greater than those who had GWG less than 6kg (p=0.027 in χ2/Fisher’s exact test). As supported by previous studies ^(5,18–20)^, gestational diabetes, and increased GWG have been reported to surge the risk of post-term births ^(5,19,20)^, which subsequently leads to relatively higher post-term births in the municipality.

This study identified some limitations. Selection bias could happen. The study did not recruit mothers without antenatal record books, delivered at home and were absent from first-trimester antenatal services. Furthermore, information bias could occur due to the use of previous obstetric and anthropometric data from antenatal records including gestational age, birthweight, birth length, ANC visits, and mother’s weight among others. This is because mismeasurement and mis-recording could occur rendering some of the data incorrect. Finally, since the study design is a cross-sectional type, it may require regional or national studies to attain generalizable findings and conclusions.

## CONCLUSION

Preterm births are very prevalent while post-term births are increasingly becoming more important in the Savelugu municipality. Preterm births were independently associated with anaemia in the first trimester of pregnancy, no ITNs used during gestation, teenage mothers, and mothers with junior high school education. Alternatively, while macrosomic deliveries increased the risk of post-term births, mothers with senior high school levels of education appeared to be a protective factor. These risk factors are modifiable and preventable. Ghanaian Ministry of Health through the Ghana Health Service should give greater attention to first-trimester anaemia during pregnancy (due to its effect on preterm births) through the consistent creation of community and health workers’ awareness. Also, promoting and encouraging girl child education through parental empowerment in Ghana (especially in Savelugu municipality) in addition to regular community education on nutrition together with lifestyle modification are commended.

## DECLARATIONS

## Ethical Standards Disclosure

This study was conducted according to the guidelines laid down in the Declaration of Helsinki and all procedures involving research study participants were approved by the Ethical Committee of Graduate School of Tropical Medicine and Global Health, Nagasaki University, Japan (approval no.: NU_TMGH_2021_194_1) and Ghana Health Service Ethics Review Committee, Ghana (approval no.: GHS-ERC 026/12/21). Written informed consent/assent was obtained from all study participants and/or legal representatives.

## Consent for publication

Not required

## Availability of data and materials

The datasets generated, and analyzed during the present study will be made available by the corresponding author, without undue reservation. Kindly

## Authorship

SAG, HA, and AA formulated the research questions, designed, and conceptualized the study. SAG and AA carried out the data collection, data analysis, and interpretations. All authors contributed to drafting the manuscript, reviewing, and approving the final manuscript.

## Authors’ acronyms

Silas Adjei-Gyamfi (SAG); Abigail Asirifi (AA); Hirotsugu Aiga (HA)

## Acknowledgements

The authors are grateful to The Project for Human Resource Development Scholarship - Japan International Cooperation Agency (JICA), Nagasaki University School of Tropical Medicine and Global Health - Japan, and Savelugu Municipal Hospital - Ghana Health Service for supporting this study. We are also thankful to all research assistants, study participants, and volunteers who contributed in diverse ways to the successful implementation of the study.

## LIST OF ABBREVIATIONS

ANC: Antenatal care
GWG: Gestational weight gain
ITNs: Insecticide-treated bed nets
MCHRBs: Maternal and child health record books
ODK: Open data kit
PNC: Postnatal care
SDGs: Sustainable development goals
SES: Socioeconomic status
SP: Sulphadoxine pyrimethamine
STATA: Statistics and data

