## Supplementary material for "Prevalence and associated risk factors of preterm and post-term births in Northern Ghana: a retrospective study in Savelugu Municipality": Supplemtental Tables 1, 2, 3, 4, 5

Table 1: Approximated facility sample sizes in the study area

| **Health facilities** | **U: Total first PNC visits at targeted health facilities in 2020** | **V: Facility coverage**  **[=(U)/8042]** | **X: Number of mothers to be drawn for each facility [= (V) x 356]** |
| --- | --- | --- | --- |
| Diare health centre | 2220 | 27.6% | 98 |
| Moglaa health centre | 346 | 4.3% | 15 |
| Pong-Tamale health centre | 852 | 10.6% | 38 |
| Savelugu health centre | 1198 | 14.9% | 53 |
| Savelugu municipal hospital | 3426 | 42.6% | 152 |
| Total | 8042 | 100.0% | 356 |

Table 2: Association of preterm and post-term births with sociodemographic variables among study participants

| **Background variables** |  |  | **Bivariate analysis** | | | |
| --- | --- | --- | --- | --- | --- | --- |
|  | **Total (%)** |  | **Preterm (%)** | **Post-term (%)** | **Normal term (%)** | **p**–**value ^*^** |
|  | **n = 356** |  | **n = 69** | **n = 23** | **n = 264** |  |
| **Maternal age category** | |  |  |  |  |  |
| 16 to 19 years | 18 (5.1) |  | 10 (14.5) | 0 (0.0) | 8 (3.0) | 0.001**^†^** |
| 20 to 35 years | 308 (86.5) |  | 54 (78.2) | 23 (100.0) | 231 (87.5) |  |
| Above 35 years | 30 (8.4) |  | 5 (7.3) | 0 (0.0) | 25 (9.5) |  |
| Mean (SD) = 27.3 (5.4) |  |  |  |  |  |  |
| **Marital status** |  |  |  |  |  |  |
| Married | 319 (89.6) |  | 59 (85.5) | 21 (91.3) | 239 (90.5) | 0.492 |
| Unmarried | 37 (10.4) |  | 10 (14.5) | 2 (8.7) | 25 (9.5) |  |
| **Highest level of education** |  |  |  |  |  |  |
| None | 121 (34.0) |  | 24 (34.8) | 13 (56.5) | 84 (31.8) | 0.009**^†^** |
| Primary | 46 (12.9) |  | 14 (20.3) | 3 (13.0) | 29 (11.0) |  |
| Junior high | 46 (12.9) |  | 7 (10.1) | 3 (13.0) | 36 (13.6) |  |
| Senior high | 105 (29.5) |  | 14 (20.3) | 1 (4.5) | 90 (34.1) |  |
| Tertiary | 38 (10.7) |  | 10 (14.5) | 3 (13.0) | 25 (9.5) |  |
| **Ethnic group** |  |  |  |  |  |  |
| Dagomba | 273 (76.7) |  | 52 (75.4) | 15 (65.2) | 206 (78.0) | 0.432 |
| Frafra | 60 (16.8) |  | 14 (20.3) | 6 (26.1) | 40 (15.2) |  |
| Others **^a^** | 23 (6.5) |  | 3 (4.4) | 2 (8.7) | 18 (6.8) |  |
| **Religious affiliation** |  |  |  |  |  |  |
| Christian | 44 (12.4) |  | 4 (5.8) | 3 (13.0) | 37 (14.0) | 0.181 |
| Muslim | 312 (87.6) |  | 65 (94.2) | 20 (87.0) | 227 (86.0) |  |
| **Occupation status** |  |  |  |  |  |  |
| Unemployed | 142 (39.9) |  | 24 (34.8) | 7 (30.4) | 111 (42.1) | 0.334 |
| Informal | 181 (50.8) |  | 40 (57.9) | 15 (65.2) | 126 (47.7) |  |
| Formal | 33 (9.3) |  | 5 (7.3) | 1 (4.4) | 27 (10.2) |  |
| **Fuel type** |  |  |  |  |  |  |
| Firewood | 148 (41.6) |  | 32 (46.4) | 7 (30.4) | 109 (41.3) | 0.397 |
| Charcoal | 162 (45.5) |  | 31 (44.9) | 14 (60.9) | 117 (44.3) |  |
| Gas | 46 (12.9) |  | 6 (8.7) | 2 (8.7) | 38 (14.4) |  |
| **Wealth index** | |  |  |  |  |  |
| Poorest | 72 (20.2) |  | 13 (18.8) | 3 (13.0) | 56 (21.2) | 0.630 |
| Poorer | 75 (21.0) |  | 19 (27.5) | 5 (21.7) | 51 (19.3) |  |
| Poor | 70 (19.7) |  | 15 (21.7) | 4 (17.4) | 51 (19.3) |  |
| Less Poor | 70 (19.7) |  | 13 (19.0) | 7 (30.4) | 50 (19.0) |  |
| Least Poor | 69 (19.4) |  | 9 (13.0) | 4 (17.4) | 56 (21.2) |  |
| **Family size** |  |  |  |  |  |  |
| < 10 occupants | 283 (79.5) |  | 58 (84.1) | 19 (82.6) | 206 (78.0) | 0.505 |
| ≥ 10 occupants | 73 (20.5) |  | 11 (15.9) | 4 (17.4) | 58 (22.0) |  |
| **Neonatal age category** |  |  |  |  |  |  |
| < 1 week | 106 (29.8) |  | 20 (29.0) | 6 (26.1) | 80 (30.3) | 0.902 |
| 2 – 4 weeks | 250 (70.2) |  | 49 (71.0) | 17 (73.9) | 184 (69.7) |  |
| **Gender of neonates** |  |  |  |  |  |  |
| Male | 200 (56.2) |  | 38 (55.1) | 10 (43.5) | 152 (57.6) | 0.417 |
| Female | 156 (43.8) |  | 31 (44.9) | 13 (56.5) | 112 (42.4) |  |

*SD: standard deviation*	***^†^*** *p-value < 0.05*	***^*^*** *Chi-square/Fisher’s exact test*	***^a^*** *Bulsa, Dagaati, Ewe*

Table 3: Association of preterm and post-term births with antenatal and anthropometric variables among study participants

| **Background variables** |  |  | **Bivariate analysis** | | | |
| --- | --- | --- | --- | --- | --- | --- |
|  | **Total (%)** |  | **Preterm (%)** | **Post-term (%)** | **Normal term (%)** | **p**–**value ^*^** |
|  | **n = 356** |  | **n = 69** | **n = 23** | **n = 264** |  |
| **Maternal body mass index (BMI)** |  |  |  |  |  |  |
| Underweight (< 18.5 kg/m^2^) | 25 (7.0) |  | 4 (5.8) | 0 (0.0) | 21 (8.0) | 0.540 |
| Normal BMI (≥ 18.5 to 24.9 kg/m^2^) | 229 (64.3) |  | 47 (68.1) | 18 (78.3) | 164 (62.1) |  |
| Overweight (≥ 25.0 kg/m^2^) | 102 (28.7) |  | 18 (26.1) | 5 (21.7) | 79 (29.9) |  |
| **First-trimester gestational weight** |  |  |  |  |  |  |
| < 50 kg | 42 (11.8) |  | 9 (13.0) | 0 (0.0) | 33 (12.5) | 0.192 |
| ≥ 50 kg | 314 (88.2) |  | 60 (87.0) | 23 (100.0) | 231 (87.5) |  |
| **Maternal height** | |  |  |  |  |  |
| < 150 cm | 13 (3.7) |  | 2 (2.9) | 0 (0.0) | 11 (4.2) | 1.000 |
| ≥ 150 cm | 343 (96.7) |  | 67 (97.1) | 23 (100.0) | 253 (95.8) |  |
| **Gestational weight gain** | |  |  |  |  |  |
| < 6 kg | 94 (26.4) |  | 19 (27.5) | 0 (0.0) | 75 (21.2) | 0.012**^†^** |
| ≥ 6 kg | 262 (73.6) |  | 50 (72.5) | 23 (100.0) | 189 (71.6) |  |
| **Neonatal birth length** | |  |  |  |  |  |
| ≤ 47.5 cm | 72 (20.2) |  | 16 (23.2) | 0 (0.0) | 56 (21.2) | 0.041**^†^** |
| > 47.5 cm | 284 (79.8) |  | 53 (76.8) | 23 (100.0) | 208 (78.8) |  |
| **Neonatal birth weight (BW)** |  |  |  |  |  |  |
| Low birth weight (< 2.5 kg) | 79 (22.2) |  | 29 (42.0) | 2 (8.7) | 48 (18.2) | < 0.001**^†^** |
| Normal BW (≥ 2.5 kg – 3.9 kg) | 246 (69.1) |  | 37 (53.6) | 11 (47.8) | 198 (75.0) |  |
| Macrosomia (≥ 4.0 kg) | 31 (8.7) |  | 3 (4.4) | 10 (43.5) | 18 (6.8) |  |
| **Frequency of ANC visits** |  |  |  |  |  |  |
| < 8 visits | 191 (53.7) |  | 48 (69.6) | 4 (17.4) | 139 (52.6) | < 0.001**^†^** |
| ≥ 8 visits | 165 (46.3) |  | 21 (30.4) | 19 (82.6) | 125 (47.4) |  |
| **Facility type for ANC visits** |  |  |  |  |  |  |
| District hospital | 175 (49.2) |  | 32 (46.4) | 10 (43.5) | 133 (50.4) | 0.716 |
| Health centre | 181 (50.8) |  | 37 (53.6) | 13 (56.5) | 131 (49.6) |  |
| **Family planning use before last pregnancy** | |  |  |  |  |  |
| No | 232 (65.2) |  | 45 (65.2) | 12 (52.2) | 175 (66.3) | 0.395 |
| Yes | 124 (34.8) |  | 24 (34.8) | 11 (47.8) | 89 (33.7) |  |
| **Insecticide-treated nets (ITNs) use** |  |  |  |  |  |  |
| No | 138 (38.8) |  | 36 (52.2) | 9 (39.1) | 93 (35.2) | 0.037**^†^** |
| Yes | 218 (61.2) |  | 33 (47.8) | 14. (60.9) | 171 (64.8) |  |
| **Frequency of sulphadoxine-pyrimethamine (SP) intake** | |  |  |  |  |  |
| None | 18 (5.1) |  | 6 (8.7) | 2 (8.7) | 10 (3.8) | 0.009**^†^** |
| 1 to 3 times | 141 (39.6) |  | 34 (49.3) | 3 (13.0) | 104 (39.4) |  |
| Above 3 times | 197 (55.3) |  | 29 (42.0) | 18 (78.3) | 150 (56.8) |  |
| **Anthelminthics intake** | |  |  |  |  |  |
| No | 218 (61.2) |  | 44 (63.8) | 15 (65.2) | 159 (60.2) | 0.797 |
| Yes | 138 (38.8) |  | 25 (36.2) | 8 (34.8) | 105 (39.8) |  |
| **Iron/folic acid intake** |  |  |  |  |  |  |
| No | 13 (3.7) |  | 1 (1.5) | 0 (0.0) | 12 (4.6) | 0.298 |
| Yes | 343 (96.3) |  | 68 (98.5) | 23 (100.0) | 252 (95.4) |  |
| **Tetanus-diphtheria vaccination** |  |  |  |  |  |  |
| No | 26 (7.3) |  | 4 (5.8) | 2 (8.7) | 20 (7.6) | 0.850 |
| Yes | 330 (92.7) |  | 65 (94.2) | 21 (91.3) | 244 (92.4) |  |
| **Nutrition education received** | |  |  |  |  |  |
| No | 47 (13.2) |  | 5 (7.3) | 6 (26.1) | 36 (13.6) | 0.064 |
| Yes | 309 (86.8) |  | 64 (92.7) | 17 (73.9) | 228 (86.4) |  |

***^†^*** *p-value < 0.05*	***^*^*** *Chi-square test/Fisher’s exact test*

Table 4: Association of preterm and post-term births with clinical and obstetric variables among study participants

| **Background variables** |  |  | **Bivariate analysis** | | | |
| --- | --- | --- | --- | --- | --- | --- |
|  | **Total (%)** |  | **Preterm (%)** | **Post-term (%)** | **Normal term (%)** | **p-value ^*^** |
|  | **n = 356** |  | **n = 69** | **n = 23** | **n = 264** |  |
| **Number of pregnancies** |  |  |  |  |  |  |
| 0 – 1 pregnancy | 88 (24.7) |  | 25 (36.2) | 0 (0.0) | 63 (23.9) | 0.002**^†^** |
| ≥ 2 pregnancies | 268 (75.3) |  | 44 (63.8) | 23 (100.0) | 201 (76.1) |  |
| **Number of deliveries** |  |  |  |  |  |  |
| 0 – 1 delivery | 97 (27.3) |  | 27 (39.1) | 0 (0.0) | 70 (26.5) | 0.001**^†^** |
| ≥ 2 deliveries | 259 (72.7) |  | 42 (60.9) | 23 (100.0) | 194 (73.5) |  |
| **Haemoglobin (Hb) levels at first trimester** | |  |  |  |  |  |
| Low Hb/Anaemia (< 11g/dL) | 160 (44.9) |  | 49 (71.0) | 4 (17.4) | 107 (40.5) | < 0.001**^†^** |
| Normal Hb (≥ 11 – 13.1g/dL) | 177 (49.7) |  | 20 (29.0) | 12 (52.2) | 145 (54.9) |  |
| High Hb (≥ 13.2g/dL) | 19 (5.4) |  | 0 (0.00) | 7 (30.4) | 12 (4.6) |  |
| **Hb levels at second trimester** | |  |  |  |  |  |
| Low Hb/Anaemia | 200 (56.2) |  | 56 (81.2) | 5 (21.7) | 139 (52.6) | < 0.001**^†^** |
| Normal Hb | 152 (42.7) |  | 13 (18.8) | 16 (69.6) | 123 (46.6) |  |
| High Hb | 4 (1.1) |  | 0 (0.00) | 2 (8.7) | 2 (0.8) |  |
| **Hb levels at third trimester** | |  |  |  |  |  |
| Low Hb/Anaemia | 158 (44.4) |  | 46 (66.6) | 3 (13.0) | 109 (41.3) | < 0.001**^†^** |
| Normal Hb | 192 (53.9) |  | 22 (31.9) | 19 (82.6) | 151 (57.2) |  |
| High Hb | 6 (1.7) |  | 1 (1.5) | 1 (4.4) | 4 (1.5) |  |
| **Malaria infection during pregnancy** | |  |  |  |  |  |
| No | 254 (71.3) |  | 44 (63.8) | 20 (87.0) | 190 (72.0) | 0.094 |
| Yes | 102 (28.7) |  | 25 (36.2) | 3 (13.0) | 74 (28.0) |  |
| **HIV infection** |  |  |  |  |  |  |
| No | 350 (98.3) |  | 67 (97.1) | 22 (95.6) | 261 (98.9) | 0.354 |
| Yes | 6 (1.7) |  | 2 (2.9) | 1 (4.4) | 3 (1.1) |  |
| **Hepatitis B infection** |  |  |  |  |  |  |
| No | 318 (89.3) |  | 58 (84.1) | 21 (91.3) | 239 (90.5) | 0.286 |
| Yes | 38 (10.7) |  | 11 (15.9) | 2 (8.7) | 25 (9.5) |  |
| **Gestational diabetes status** |  |  |  |  |  |  |
| Not diagnosed | 335 (94.1) |  | 69 (100.0) | 20 (87.0) | 246 (93.2) | 0.014**^†^** |
| Diagnosed | 21 (5.9) |  | 0 (0.00) | 3 (13.0) | 18 (6.8) |  |
| **Gestational hypertension status** |  |  |  |  |  |  |
| Not diagnosed | 312 (87.6) |  | 58 (86.1) | 20 (87.0) | 234 (88.6) | 0.586 |
| Diagnosed | 44 (12.4) |  | 11 (15.9) | 3 (13.0) | 30 (11.4) |  |
| **Sickle cell status** |  |  |  |  |  |  |
| Negative | 301 (84.5) |  | 54 (78.3) | 18 (78.3) | 229 (86.7) | 0.153 |
| Positive | 55 (15.5) |  | 15 (21.7) | 5 (21.7) | 35 (13.3) |  |
| **G6PD status** |  |  |  |  |  |  |
| Normal | 333 (93.5) |  | 61 (88.4) | 20 (87.0) | 252 (95.4) | 0.034**^†^** |
| Complete/partial | 23 (6.5) |  | 8 (11.6) | 3 (13.0) | 12 (4.6) |  |

***^†^*** *p-value < 0.05*	***^*^*** *Chi-square/Fisher’s exact test*

**Table 5: Multivariate analysis of risk factors for preterm and post-term births**

|  | **Multinomial logistic regression *(Normal term at delivery = base outcome)*** | | | | | | | | |
| --- | --- | --- | --- | --- | --- | --- | --- | --- | --- |
|  | **Preterm** | | | |  | **Post-term** | | | |
| **Variables** | **COR** | **p**–**value (95%CI)** | **AOR** | **p**–**value (95%CI)** |  | **COR** | **p**–**value (95%CI)** | **AOR** | **p**–**value (95%CI)** |
| **Maternal age category** |  |  |  |  |  |  |  |  |  |
| 16 to 19 years | 5.344 | 0.001 (2.014 – 14.18)**^†^** | **12.95** | **0.001 (2.977 – 56.34)^†^** |  | ––– | 0.994 (–––) | ––– | 0.997 (–––) |
| 20 to 35 years |  |  |  | 1.00 |  |  |  |  | 1.00 |
| Above 35 years | 0.856 | 0.761 (0.313 – 2.337) | 0.651 | 0.482 (0.197 – 2.154) |  | ––– | 0.989 (–––) | ––– | 0.993 (–––) |
| **Highest level of education** | |  |  |  |  |  |  |  |  |
| None |  |  |  | 1.00 |  |  |  |  | 1.00 |
| Primary school | 1.690 | 0.189 (0.772 – 3.696) | 1.077 | 0.878 (0.418 – 2.770) |  | 0.668 | 0.551 (0.178 – 2.513) | 0.176 | 0.079 (0.252 – 1.227) |
| Junior high school | 0.681 | 0.016 (0.269 – 1.722)**^†^** | **0.225** | **0.017 (0.065** – **0.797)^†^** |  | 0.538 | 0.356 (0.145 – 2.005) | 0.237 | 0.152 (0.033 – 1.704) |
| Senior high school | 0.544 | 0.099 (0.264 – 1.122) | 0.382 | 0.057 (0.154 – 0.944) |  | 0.072 | 0.012 (0.009 – 0.561)**^†^** | **0.001** | **0.004 (0.0001 – 0.125)^†^** |
| Tertiary | 1.400 | 0.444 (0.591 – 3.316) | 1.282 | 0.647 (0.442 – 3.717) |  | 0.775 | 0.708 (0.205 – 2.939) | 0.111 | 0.068 (0.010 – 1.179) |
| **Gestational weight gain** |  |  |  |  |  |  |  |  |  |
| < 6 kg |  |  |  | 1.00 |  |  |  |  | 1.00 |
| ≥ 6 kg | 1.044 | 0.006 (0.578 – 1.888)**^†^** | 1.754 | 0.123 (0.860 – 3.578) |  | ––– | 0.991 (–––) | ––– | 0.991 (–––) |
| **Neonatal birth length** |  |  |  |  |  |  |  |  |  |
| ≤ 47.5 cm |  |  |  | 1.00 |  |  | 1.00 |  | 1.00 |
| > 47.5 cm | 0.891 | 0.022 (0.474 – 1.678)**^†^** | 2.039 | 0.079 (0.919 – 4.521) |  | ––– | 0.974 (–––) | ––– | 0.992 (–––) |
| **Frequency of ANC visits** |  |  |  |  |  |  |  |  |  |
| < 8 visits |  |  |  | 1.00 |  |  |  |  | 1.00 |
| ≥ 8 visits | 0.487 | 0.013 (0.276 – 0.858)**^†^** | 0.862 | 0.688 (0.417 – 1.780) |  | 5.282 | 0.003 (1.750 – 15.95)**^†^** | 4.652 | 0.111 (0.702 – 30.82) |
| **ITNs use** |  |  |  |  |  |  |  |  |  |
| Yes |  |  |  | 1.00 |  |  |  |  | 1.00 |
| No | 2.006 | 0.011 (1.174 – 3.427)**^†^** | **1.979** | **0.005 (0.999 – 3.920)^†^** |  | 1.182 | 0.708 (0.493 – 2.835) | 1.244 | 0.772 (0.284 – 5.442) |
| **Frequency of SP intake** |  |  |  |  |  |  |  |  |  |
| None | 1.835 | 0.272 (0.621 – 5.424) | 0.529 | 0.596 (0.050 – 5.594) |  | 6.933 | 0.046 (1.034 – 46.51)**^†^** | 5.434 | 0.112 (0.393 – 7.520) |
| 1 to 3 times |  |  |  | 1.00 |  |  |  |  | 1.00 |
| Above 3 times | 0.591 | 0.064 (0.339 – 1.030) | 0.894 | 0.746 (0.452 – 1.767) |  | 4.160 | 0.025 (1.195 – 14.48)**^†^** | 4.641 | 0.088 (0.794 – 27.13) |
| **Number of pregnancies** |  |  |  |  |  |  |  |  |  |
| 0 – 1 pregnancy |  | [Reference] | Excluded due to multicollinearity | |  |  | [Reference] | Excluded due to multicollinearity | |
| ≥ 2 pregnancies | 1.812 | 0.040 (1.028 – 3.193)**^†^** |  |  |  | ––– | 0.986 (–––) |  |  |
| **Number of deliveries** |  |  |  |  |  |  |  |  |  |
| 0 – 1 delivery |  |  |  | 1.00 |  |  |  |  | 1.00 |
| ≥ 2 deliveries | 1.781 | 0.042 (1.022 – 3.104)**^†^** | 1.005 | 0.989 (0.451 – 2.242) |  | ––– | 0.987 (–––) | ––– | 0.991 (–––) |
| **Neonatal birth weight** |  |  |  |  |  |  |  |  |  |
| Low birth weight | 3.233 | < 0.001 (1.811 – 5.771)**^†^** | 1.514 | 0.337 (0.649 – 3.535) |  | 0.750 | 0.714 (0.161 – 3.496) | 1.270 | 0.864 (0.083 – 19.39) |
| Normal BW |  |  |  | 1.00 |  |  |  |  | 1.00 |
| Macrosomia | 0.892 | 0.860 (0.250 – 3.181) | 2.845 | 0.172 (0.634 – 12.77) |  | 10.00 | < 0.001 (3.742 – 26.72)**^†^** | **8.128** | **0.007 (1.777 – 37.18)^†^** |
| **Hb levels at first trimester** | |  |  |  |  |  |  |  |  |
| Low Hb/Anaemia | 3.320 | < 0.001 (1.865 – 5.911)**^†^** | **2.205** | **0.047 (1.011 – 4.809)^†^** |  | 0.452 | 0.179 (0.142 – 1.439) | 1.270 | 0.864 (0.083 – 19.39) |
| Normal Hb |  |  |  | 1.00 |  |  |  |  | 1.00 |
| High Hb | ––– | 0.991 (–––) | ––– | 0.998 (–––) |  | 7.051 | 0.001 (2.341 – 21.23)**^†^** | 4.716 | 0.162 (0.538 – 41.35) |
| **Hb levels at second trimester** | |  |  |  |  |  |  |  |  |
| Low Hb/Anaemia | 3.812 | < 0.001(1.989 – 7.304)**^†^** | 2.422 | 0.050 (0.998 – 5.876) |  | 0.277 | 0.015 (0.098 – 0.777)**^†^** | 0.747 | 0.726 (0.147 – 3.798) |
| Normal Hb |  |  |  | 1.00 |  |  |  |  | 1.00 |
| High Hb | ––– | 0.987 (–––) | ––– | 0.999 (–––) |  | 7.944 | 0.049 (1.010 – 58.34)**^†^** | 0.136 | 0.236 (0.005 – 3.658) |
| **Hb levels at third trimester** | |  |  |  |  |  |  |  |  |
| Low Hb/Anaemia | 2.897 | < 0.001 (1.647 – 5.094)**^†^** | 1.009 | 0.983 (0.449 – 2.268) |  | 0.219 | 0.016 (0.063 – 0.757)**^†^** | 0.427 | 0.440 (0.049 – 3.699) |
| Normal Hb |  |  |  | 1.00 |  |  |  |  | 1.00 |
| High Hb | 1.715 | 0.636 (0.183 – 16.06) | 14.46 | 0.097 (0.614 – 34.07) |  | 1.987 | 0.549 (0.211 – 18.71) | 0.288 | 0.458 (0.011 – 7.743) |
| **Gestational diabetes status** | |  |  |  |  |  |  |  |  |
| Not diagnosed |  |  |  | 1.00 |  |  |  |  | 1.00 |
| Diagnosed | ––– | 0.982 (–––) | ––– | 0.998 (–––) |  | 2.051 | 0.028 (0.557 – 7.558)**^†^** | 3.814 | 0.221 (0.448 – 32.47) |
| **G6PD status** |  |  |  |  |  |  |  |  |  |
| Normal |  |  |  | 1.00 |  |  |  |  | 1.00 |
| Complete/partial | 2.754 | 0.034 (1.078 – 7.031)**^†^** | 2.673 | 0.337 (0.360 – 19.87) |  | 3.150 | 0.094 (0.821 – 12.08) | 6.242 | 0.285 (0.218 – 17.89) |
| ***Regression model*** |  |  |  |  |  |  |  |  |  |
| *R^2^* | *0.332* |  |  |  |  |  |  |  |  |
| *p-value* | *< 0.001* |  |  |  |  |  |  |  |  |

***^†^*** *p-value < 0.05	AOR: Adjusted odds ratio	COR: Crude odds ratio*
